# Genome-wide association studies of cognitive and motor progression in Parkinson’s disease

**DOI:** 10.1101/2020.05.07.20084343

**Authors:** Manuela MX Tan, Michael A Lawton, Edwin Jabbari, Regina H Reynolds, Hirotaka Iwaki, Cornelis Blauwendraat, Sofia Kanavou, Miriam I Pollard, Leon Hubbard, Naveed Malek, Katherine A Grosset, Sarah L Marrinan, Nin Bajaj, Roger A Barker, David J Burn, Catherine Bresner, Thomas Foltynie, Nicholas W Wood, Caroline H Williams-Gray, John Hardy, Michael A Nalls, Andrew B Singleton, Nigel M Williams, Yoav Ben-Shlomo, Michele TM Hu, Donald G Grosset, Maryam Shoai, Huw R Morris

## Abstract

**Background:** There are currently no treatments that stop or slow the progression of Parkinson’s disease (PD). Case-control genome-wide association studies (GWASs) have identified variants associated with disease risk, but not progression.

**Objective:** To identify genetic variants associated with PD progression in GWASs.

**Methods:** We analysed three large, longitudinal cohorts: Tracking Parkinson’s, Oxford Discovery, and the Parkinson’s Progression Markers Initiative. We included clinical data for 3,364 patients with 12,144 observations (mean follow-up 4.2 years). We used a new method in PD, following a similar approach in Huntington’s disease, where we combined multiple assessments using a principal components analysis to derive scores for composite, motor, and cognitive progression. These scores were analysed in linear regressions in GWASs. We also performed a targeted analysis of the 90 PD risk loci from the latest case-control meta-analysis.

**Results:** There was no overlap between variants associated with PD risk, from case-control studies, and PD age at onset versus PD progression. The *APOE* ε4 tagging variant, rs429358, was significantly associated with the rate of composite and cognitive progression in PD. No single variants were associated with motor progression. However in gene-based analysis, variation across *ATP8B2*, a phospholipid transporter related to vesicle formation, was nominally associated with motor progression (p=5.3 × 10^−6^).

**Conclusions:** This new method in PD improves measurement of symptom progression. We provide strong evidence that the *APOE* ε4 allele drives progressive cognitive impairment in PD. We have also identified other loci of interest, such as *ATP8B2* for motor progression, which need to be replicated in further studies.

## INTRODUCTION

Progression in Parkinson’s disease (PD) is heterogeneous, with some patients progressing rapidly while others remain relatively stable over time^1^. There is a clear need to identify genetic variants that affect symptom progression in PD. These genes and pathways could be targeted to develop therapies to stop or slow progression of PD. Genetic factors could also help to stratify patients and predict progression more accurately in clinical trials.

Genome-wide association studies (GWASs) in PD have identified 90 independent loci associated with disease risk^2^. However, the majority of PD GWASs have compared cases to healthy controls to identify variants linked to disease status. In order to identify variants that are associated with disease progression, it is necessary to compare phenotypes within patients.

Progression of clinical signs in PD can be measured in different ways^3^ and there is no gold standard measure of progression, although the MDS-UPDRS Part III and Part II are commonly used in clinical trials. Individual scales, including the MDS-UPDRS, are affected by measurement error particularly for change over time^4^, including rater subjectivity and practice effects in cognitive assessments. Therefore, combining multiple measures may improve the accuracy of measuring progression^5,6^, as shown in the Huntington’s disease (HD) progression GWAS^7^. In this study, we analysed data from three large-scale, prospective, longitudinal studies: Tracking Parkinson’s, Oxford Parkinson’s Disease Centre Discovery, and Parkinson’s Progression Markers Initiative (PPMI). We combined multiple measures of motor and cognitive progression using Principal Components Analysis (PCA) to create progression scores. These scores were analysed in GWASs to identify variants associated with composite (crossdomain), motor, and cognitive progression in PD.

## METHODS

### Cohorts

Tracking Parkinson’s is an observational, UK multi-centre study^8^. Participants with a clinical diagnosis of PD were recruited between 2012 and 2014. Standardised clinical assessments were conducted every 1.5 years. Ethical approval was granted by the West of Scotland Research Ethics Service Research Ethics Committee.

Oxford Discovery is another UK observational, multi-centre study^9^. The inclusion criteria were the same as in Tracking Parkinson’s, and almost the same assessments were collected every 1.5 years. Ethical approval was granted by the Berkshire Research Ethics Committee.

PPMI (http://www.ppmi-info.org/) is a multi-centre study of newly diagnosed PD patients^10^. PPMI data was downloaded on 14/08/2019. Only data from the annual visits was analysed from PPMI, as the motor assessments were performed in the technically defined ‘off’ state at these visits. Ethical approval was granted by local ethics committees at the participating sites.

All studies used the same Queen Square Brain Bank diagnostic criteria for PD. Patients who received alternative diagnoses during follow-up or had neuroimaging results conflicting with a PD diagnosis were excluded from analyses.

### Genotyping

DNA samples from Tracking Parkinson’s were genotyped using the Illumina HumanCore Exome array with custom content^8^. Samples from Oxford Discovery were genotyped on the Illumina HumanCore Exome-12v1.1 or the Illumina InfiniumCore Exome-24 v1.1 arrays. For PPMI, whole-genome sequencing data was used (see https://www.ppmi-info.org/). Only variants that passed filters in the joint calling process were included.

Standard quality control procedures were performed in PLINK v1.9. The cohorts were genotyped and filtered separately, but following the same quality control steps. Individuals with low overall genotyping rates (<98%), related individuals (Identity-By-Descent PIHAT>0.1), and heterozygosity outliers (>2SDs away from the mean) were removed, as were individuals whose clinically reported biological sex did not match genetically determined sex.

PCA was conducted on a linkage disequilibrium (LD) pruned set of variants (removing SNPs with an r^2^ >0.05 in a 50kb sliding window shifting 5 SNPs at a time) after merging with European samples from the HapMap reference panel. Individuals who were >6SDs away from the mean of any of the first 10 principal components were removed.

Variants were removed if they had a low genotyping rate (<99%), Hardy-Weinberg Equilibrium p-value < 1 × 10^−5^ and minor allele frequency < 1%.

Following quality control, genotypes for Tracking Parkinson’s and Oxford Discovery were imputed separately to the 1,000 Genomes Project reference panel (phase 3 release 5)^11^ using the Michigan Imputation Server (https://imputationserver.sph.umich.edu). Only variants with imputation quality >0.8 were retained, to keep only high quality calls to merge across the cohorts. Tracking Parkinson’s and Oxford Discovery data was lifted over to genome build hg38 using liftOver (https://genome.ucsc.edu/cgi-bin/hgLiftOver) to merge with PPMI. PPMI data was not imputed as this was whole-genome sequencing data.

The three datasets were then merged, with only shared variants retained. Twenty genetic principal components were generated from a linkage-pruned SNP set (removing SNPs with an r^2^ >0.02 in a 1000kb sliding window shifting 10 SNPs at a time). The first 2 components were plotted to check that there were no differences between the cohorts. We removed extreme outliers from the first 5 principal components (>6SDs away from the mean). The genetic principal components were then recalculated after removing outliers. These first 5 new principal components were included as covariates in the GWAS to adjust for population substructure. Additional outliers who were >6SDs away from the mean of any of the first 5 principal components were excluded. The plot of the first 2 principal components after removing outliers is shown in Supplementary Figures 1–2.

### Clinical outcome measures

Individual-level data from the cohorts was merged. In order to increase power and the accuracy of the final progression scores, we performed all transformations and created progression scores from the merged dataset as follows (Figure 1).

**Figure 1.**
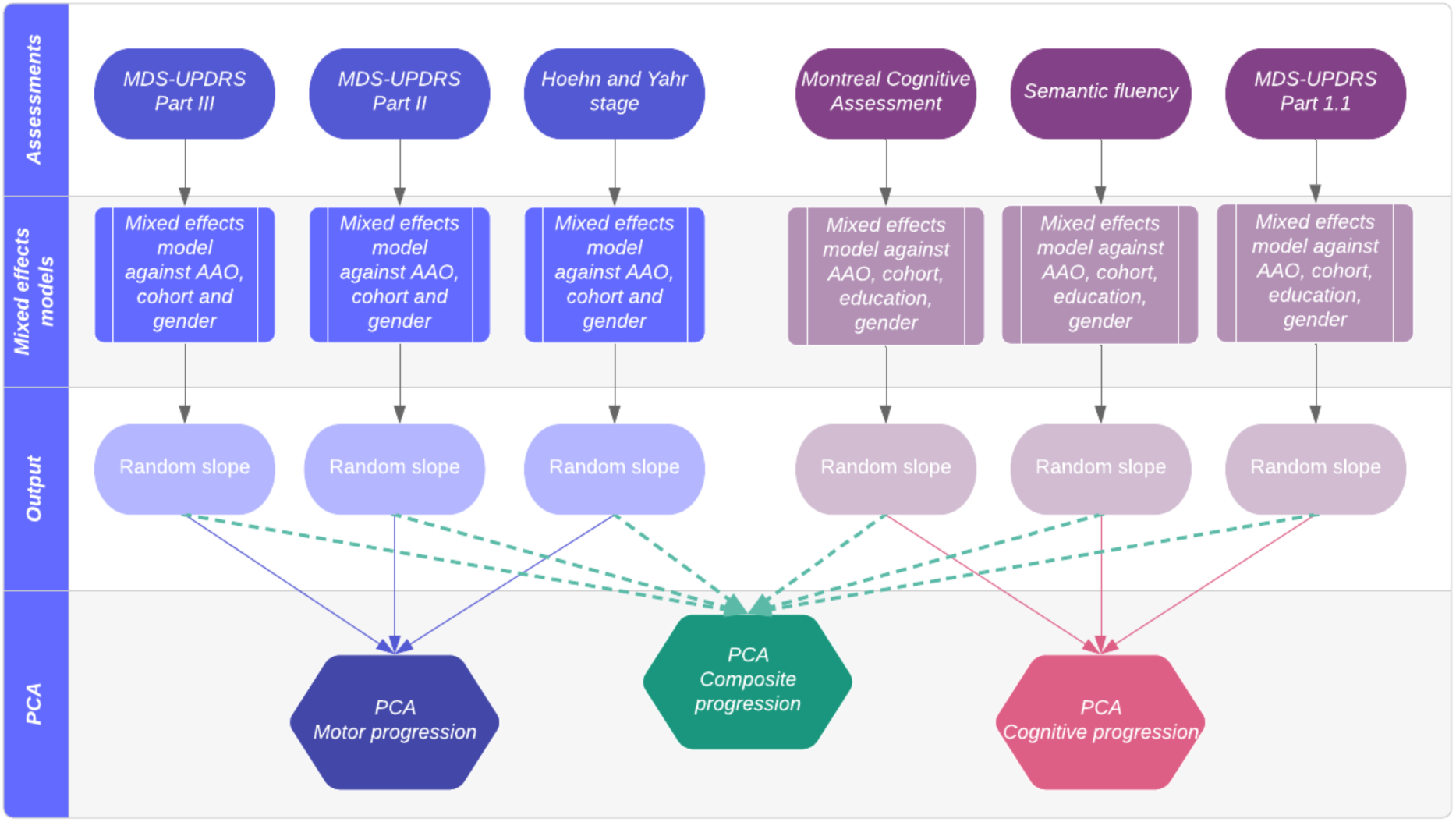
Steps to create composite, motor, and cognitive progression scores.

The motor and cognitive measures were chosen prior to the analysis. Only assessments conducted in all cohorts were included. We selected measures shown to rate motor and cognitive function semi-objectively, in an attempt to minimise observer bias. We did not include scales which may be affected by a combination of motor, cognitive, and other non-motor symptoms.

Motor progression was assessed using the MDS-UPDRS Part III (clinician-assessed movement examination), MDS-UPDRS Part II (patient-reported experiences of daily living), and Hoehn and Yahr stage (clinician-assessed rating of impairment and disability)^12,13^. In PPMI, we used motor assessments conducted in the ‘off’ medication state.

Cognitive progression was assessed using the Montreal Cognitive Assessment (MoCA), semantic fluency, and item 1.1 of the MDS-UPDRS (cognitive impairment based on patient and/or caregiver report).

To ensure the different measures were comparable, we first converted raw scores into a percentage of the maximum score for that scale, with higher scores indicating worse symptoms. The MoCA was reverse scored to count the number of incorrect items out of the maximum score of 30. Semantic fluency was reverse scored out of the highest individual score at baseline in each cohort.

Each measure was normalised to the population baseline mean and standard deviation within each cohort, to adjust for any differences in the scales or task instructions between cohorts. This also ensures that the measures are on the same scale, and preserves data on longitudinal change.

### Analysis

#### Progression scores

We derived severity scores from mixed effects regression models using follow-up data up to 72 months. Each variable was regressed on age at onset, sex, cohort, and their interactions with time from disease onset. For the cognitive measures, we also included the number of years of education before higher education, and whether higher education was undertaken (yes/no). In each model, we included terms for subject random effects to account for individual heterogeneity in the intercept (baseline values) and slope (rate of progression).

We used the random effect slope values as the measure of ‘residual’ progression not predicted by age at onset, cohort, gender, and education, for each individual. We performed PCA on these values, with the input variables zero centred and scaled to have unit variance.

#### Removal of non-PD cases

Any patients that were diagnosed with a different condition during follow-up were removed from analyses. We also conducted sensitivity analyses to remove any cases which may have non-PD conditions but an alternative diagnosis had not yet been confirmed. Firstly, we repeated analyses after removing patients in Tracking Parkinson’s and Oxford Discovery who had a clinician-rated diagnostic certainty of PD <90%^14,15^. Secondly, we removed the fastest and slowest progressors in the top and bottom 5% of the distribution, to address the possibility of confounding by misdiagnosis with more benign (e.g. essential tremor) or more malignant (e.g. multiple system atrophy) conditions.

#### GWAS

Statistical analysis was conducted in R v3.4.1^16^. Clinical data was managed and cleaned using STATA (version 15.1, StataCorp, Texas, USA).

For each GWAS, we included the following covariates: cohort (to adjust for differences in genotyping data and measurement error) and the first 5 genetic principal components generated from the merged genotype data (to adjust for population substructure). GWASs were conducted in rvtests^17^ using the single variant Wald test. Genome-wide Complex Trait Analysis conditional and joint analysis (GCTA-COJO) was used to identify independent signals^18,19^.

Functional Mapping and Annotation of GWAS (FUMA; https://fuma.ctglab.nl/) was used with standard settings to annotate, prioritise, and visualize GWAS results^20^. Gene-based and gene-set analyses were conducted in FUMA with Multi-marker Analysis of GenoMic Annotation (MAGMA). MAGMA maps SNPs to genes and then tests the association between the genes and phenotype (gene-based analysis)^21^. We looked for enrichment of gene-sets or pathways in Gene Ontology (GO; MsigDB c5), Reactome, and Kyoto Encyclopedia of Genes and Genomes (KEGG). GTEx (https://gtexportal.org/) and the eQTLGen Consortium (http://www.eqtlgen.org/index.html) were used to look up expression quantitative trait loci (eQTLs). LDlink (https://ldlink.nci.nih.gov/) was used to calculate linkage between SNP pairs (using LDpair) in European populations excluding the Finnish population.

Genetic risk scores were calculated from the 90 loci from the PD case-control GWAS^2^. If SNPs were missing in our genotype data, proxies were identified using LDproxy if r^2^ >0.9. The association between the genetic risk score and each progression score was assessed using linear regression, adjusting for cohort and the first 5 genetic principal components. LD Score regression (LDSC)^22,23^ was used to estimate the genetic correlation between the progression GWASs and the PD case-control GWAS using summary statistics excluding 23andMe samples^2^.

## RESULTS

We included clinical data for 3,364 PD patients with 12,144 observations (Table 1). The mean follow-up time was 4.2 years (SD=1.5 years), and mean disease duration at study entry 2.9 years (SD=2.6 years). 79.7% of patients had completed the 72-month follow-up visit.

**Table 1.** Cohort demographics at baseline. Means (SD) are shown unless otherwise indicated.

| **Demographics at baseline** | **Tracking Parkinson's** | **Oxford Discovery** | **PPMI** |
| --- | --- | --- | --- |
| Number of PD patients | 1966 | 985 | 413 |
| Total number of visits analysed | 5980 | 3137 | 3066 |
| Mean length of follow-up (years) | 3.8 (1.4) | 4.3 (1.7) | 5.4 (1.2) |
| Male (%) | 65.2% | 64.2% | 65.4% |
| Age at onset (years) | 64.4 (9.8) | 64.5 (9.8) | 59.5 (10.0) |
| Age at diagnosis (years) | 66.3 (9.3) | 66.1 (9.6) | 61.0 (9.7) |
| Age at study entry (years) | 67.6 (9.3) | 67.4 (9.6) | 61.5 (9.8) |
| Disease duration - time from symptom onset to assessment (years) | 3.2 (3.0) | 2.9 (1.9) | 2.0 (2.0) |
| Time from diagnosis to assessment (years) | 1.3 (0.9) | 1.3 (0.9) | 0.5 (0.5) |
| MDS-UPDRS Part III | 22.9 (12.3) | 26.8 (11.1) | 20.7 (8.8) |
| MDS-UPDRS Part II | 9.9 (6.6) | 8.9 (6.2) | 5.8 (4.1) |
| Hoehn and Yahr stage mean* | 1.8 (0.6) | 1.9 (0.6) | 1.6 (0.5) |
| Hoehn and Yahr stage proportions* |  |  |  |
| 0 to 1.5 (%) | 48.1% | 23.2% | 44.8% |
| 2 to 2.5 (%) | 45.1% | 68.8% | 54.7% |
| 3+ (%) | 6.8% | 8.1% | 0.5% |
| MoCA total (adjusted for education) | 24.9 (3.6) | 24.5 (3.5) | 27.1 (2.3) |
| Semantic fluency<sup>†</sup> | 21.8 (6.9) | 34.7 (9.0) | 21.0 (5.4) |
| MDS-UPDRS Part I.1 | 0.5 (0.7) | 0.5 (0.6) | 0.3 (0.5) |

\* Tracking Parkinson's used the modified Hoehn and Yahr stage scale, while Oxford Discovery and PPMI used the original scale. Hoehn and Yahr stage proportions are shown as a total of the number of people with non-missing Hoehn and Yahr ratings at baseline.
<sup>+</sup> Instructions and timing for the semantic fluency task was slightly different between cohorts (completed within 60 seconds or 90 seconds). To account for these differences, we standardised all scales within each cohort separately (see Methods).

Within the motor progression PCA, using the MDS-UPDRS Part III, Part II and Hoehn and Yahr Stage, the first principal component explained 61.0% of the total variance. Within the cognitive domain PCA, using the MoCA, semantic fluency, and MDS-UPDRS 1.1, the first principal component explained 59.8% of the total variance (Supplementary Figures 3–6).

We found that the first principal components for motor and cognitive progression were moderately correlated (r=-0.35, p < 2.2 × 10^−16^; Supplementary Table 1). We therefore conducted a PCA combining all motor and cognitive measures, to create a composite progression score. The first principal component from this cross-domain PCA accounted for 41.0% of the joint variance (Supplementary Figure 7). Supplementary Tables 2–3 show the relationships between the raw scales and the final composite progression score. None of the composite, motor, or cognitive principal components were associated with cohort (all p-values >0.9).

### GWAS of composite progression

After quality control, imputation, and merging, 5,918,868 variants were available for analysis. 2,755 PD patients had composite progression scores and passed genetic quality control. The GWAS lambda was 1.02. One variant rs429358 in Chromosome 19 passed genome-wide significance (p=1.2×10^−8^, Figure 2, Supplementary Table 4, Supplementary Figures 8–9). This variant tags the *APOE* ε4 allele. In the gene-based test, *APOE, TOMM40* and *APOC1* reached significance (p<2.8×10^−6^, correcting for the number of mapped protein coding genes). The Reactome pathway cytosolic sulfonation of small molecules pathway was significantly enriched (p=6.9×10^−6^).

**Figure 2.**
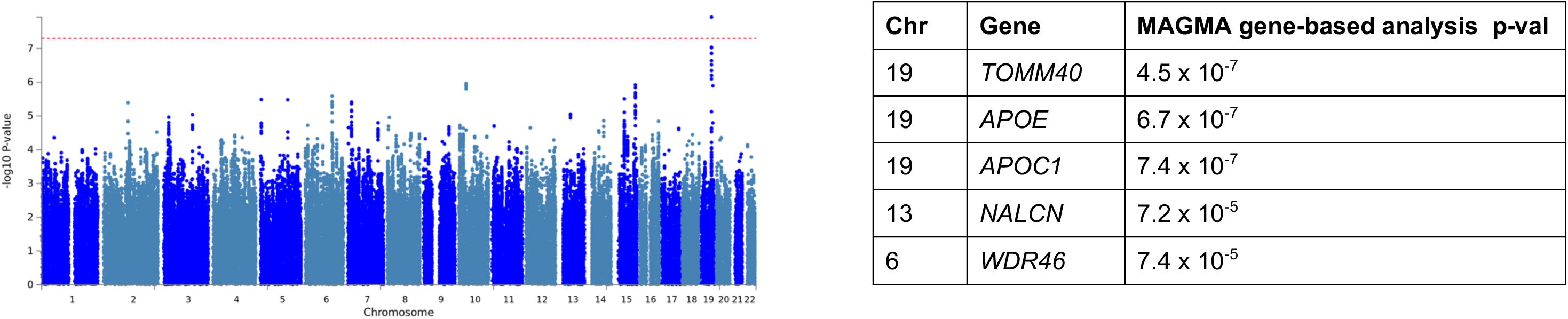
Manhattan plot for GWAS of composite progression. The red dashed line indicates the genome-wide significance threshold p-value 5 × 10^−8^.

### GWAS of motor progression

2,848 PD patients had motor progression scores and genotype data. The lambda was 1.02. No variants passed genome-wide significance (Figure 3, Supplementary Table 5). However, in the gene-based test, *ATP8B2* in Chromosome 1 was associated with motor progression (p=5.3×10^−6^, Supplementary Figures 10–11), although this did not reach significance correcting for the number of mapped genes (p=2.81×10^−6^). There was no enrichment of any gene sets or pathways.

**Figure 3.**
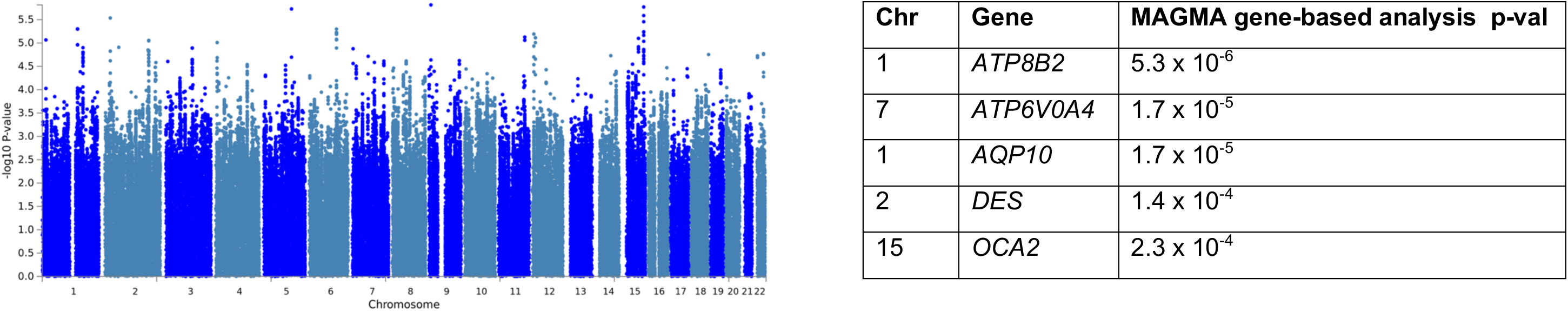
Manhattan plot for the GWAS of motor progression. Genome-wide significance is the standard p-value 5 × 10^−8^ (not indicated in the figure).

We performed follow-up analyses to confirm that the results in the top SNPs were not driven by a single cohort, or a single scale. We conducted GWASs in each cohort separately (Supplementary Table 6) and each motor scale separately (without combining in PCA). These results show that associations are strengthened with the PCA approach (Supplementary Table 7).

Our top variant in Chromosome 1, rs35950207, was associated with motor progression, p=5.0×10^−6^. We examined the associations for our top SNPs in the previous progression GWAS^24^ (https://pdgenetics.shinyapps.io/pdprogmetagwasbrowser/); rs35950207 was not significantly associated with binomial trait analysis of Hoehn and Yahr stage 3 or more at baseline (beta=0.27, p=0.03).

rs35950207 is a variant 2kb upstream of *AQP10*. It is an eQTL for *AQP10* in whole blood (GTEx p=1.7×10^−6^, eQTLGen p=3.62×10^−139^) and other tissues (subcutaneous adispose, skin, esophagus, testis, and heart). It is also an eQTL for *ATP8B2* in blood (GTEx p=1.5×10^−5^, eQTLGen p=7.84×10^−42^) and in the cerebellum (GTEx p=7.8×10^−5^). *GBA* is also located in Chromosome 1 and *GBA* variants are associated with both PD risk and progression^25^. However, rs35950207 is not in linkage disequilibrium with any of the main *GBA* variants that are implicated in PD (p.E326K, p.N370S, p.L444P, p.T369M).

rs17367669 in Chromosome 5 was the top SNP in the variant-based analysis, but there were no genes in this region that approached significance in the gene-based analysis. This variant is in an intergenic region and is closest to *LOC100505841*, Zinc Finger Protein 474-Like gene. No significant eQTLs were identified for this variant.

### GWAS of cognitive progression

2,788 patients had cognitive progression scores and genotype data. The lambda value was 1.02. The top variant was rs429358, which tags the *APOE* ε4 allele (p=2.53×10^−13^, Figure 4, Supplementary Table 8, Supplementary Figure 12–14). *APOE* was also significantly associated with cognitive progression in the gene-based analysis, in addition to *APOC1* and *TOMM40*. There was no enrichment of gene-sets or pathways. Follow-up analyses showed that the effects for the top 5 independent SNPs were consistent in each cohort and each scale (Supplementary Tables 9–10).

**Figure 4.**
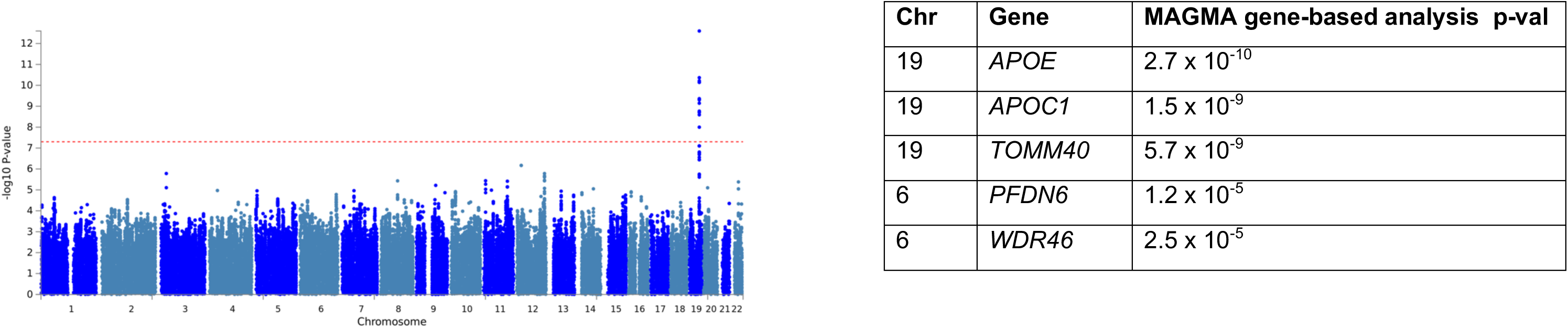
Manhattan plot for the variant-based GWAS of cognitive progression. The red dashed line indicates the genome-wide significance threshold p-value 5 × 10^−8^.

#### Targeted assessment of PD risk loci

Of the 90 risk variants from the PD case-control GWAS^2^, 73 were present in our final dataset, including the *SNCA* and *TMEM175/GAK* variants associated with PD age at onset^26^. We extracted results for these variants from the composite, motor, and cognitive GWASs. No variants passed analysis-wide significance (p=0.05/73). Variants with at least one association p<0.05 are shown in Supplementary Figure 15.

We found that only a small number of risk variants were associated with progression with p-values <0.05. rs35749011 was associated with both composite progression (beta=0.40, p=0.003) and cognitive progression (beta=-0.37, p=0.002), but not motor progression (beta=0.20, p=0.09). This variant is in linkage disequilibrium with the *GBA* p.E326K variant (also known as p.E365K), D’=0.90, R^2^=0.78.

We also extracted results for other candidate variants that have been implicated in PD progression (Supplementary Figure 16). We did not find that the top variant rs382940 in *SLC44A1* that was associated in progression to H&Y stage 3 from the Iwaki GWAS^24^ was associated with either composite, motor or cognitive progression in our GWASs (all p-values >0.05).

Overall, we did not find any overlap between the variants associated with PD risk, age at onset, and progression. Our LDSC results were also suggestive of very little overlap between the each of the progression GWASs and PD case-control GWAS (all p-values >0.5).

#### PD Genetic risk score

73 PD risk SNPs were present in our genotype data, and 2 proxies were identified for missing variants (Supplementary Table 11). There was no association between the risk score and composite progression (beta=0.09, p=0.10), or motor progression (beta=0.02, p=0.65). There was a small association between the genetic risk score and cognitive progression (beta=-0.099, p=0.04).

#### Removal of non-PD cases

We conducted sensitivity analyses to remove patients with potential non-PD conditions. Removing patients with <90% diagnostic certainty did not substantially affect our results; the top signals had slightly weaker associations in these sensitivity analyses. When we removed the extreme 5% of progressors, the top results from the main GWASs had larger p-values, although the direction of effects were the same (Supplementary Tables 12–13).

## DISCUSSION

We used a new method of analysing clinical progression in PD, by combining multiple assessments in a data-driven PCA to derive scores of composite, motor, and cognitive progression.

Our study contributes to evidence that improving the phenotypic measure can increase power in genetic studies. We showed that associations at the top signals strengthened when using the combined motor and cognitive progression scores compared to using the scales separately. The HD progression GWAS also showed that motor, cognitive, and brain imaging measures were well correlated and successfully identified a variant in *MSH3* associated with composite progression^7^. Other studies have shown that prediction accuracy of PD status or progression (such as development of cognitive impairment) is improved by combining multiple clinical, genetic, and biomarker factors^6,27^.

In PD, there are many different scales for assessing symptoms. Each scale has a degree of measurement error^4^ and different sensitivity to progression of underlying symptoms^28^. PCA is a data-driven approach that combines multiple measures to identify latent components that explain the most variability in the data, and these may more accurately reflect disease progression.

Our progression GWASs have identified two positive findings. Firstly, we replicated previous findings for *APOE* ε4. Many studies have shown that the ε4 allele is associated with more dementia in PD^29–32^, and potentially separately from the risk of Alzheimer’s disease (AD)^33^. One possible mechanism is that *APOE* is associated with amyloid-β pathology, as comorbid AD pathology is common in PD patients with dementia (PDD) at postmortem^34^. Alternatively, *APOE* may drive cognitive decline independently of amyloid/AD pathology. Recent animal model work has shown that the ε4 allele is independently associated with α-synuclein pathology and toxicity^35^. In addition, the ε4 allele is overrepresented in Dementia with Lewy Body cases with ‘pure’ Lewy body pathology, compared to PDD cases^36^. A systematic review showed that limbic and neocortical α-synuclein pathology had the strongest association with dementia in PD^34^. Further work is needed to determine the mechanisms by which *APOE* influences cognitive decline.

We identified a novel signal in *ATP8B2* associated with motor progression in a gene-based analysis. This gene encodes an ATPase phospholipid transporter (type 8B, member 2). Phospholipid translocation may be important in the formation of transport vesicles^37^. This gene has not previously been reported in PD or other diseases, and needs to be tested in other independent cohorts.

We have shown that the genetics of PD risk and progression are largely separate. We performed a targeted analysis of the 90 risk loci identified in PD case-control GWAS^2^. *GBA* p.E326K was nominally associated with composite and cognitive progression. Previous studies show that *GBA* variants are associated with rapid progression and mortality^38–43^, however many of these studies have longer follow-up, or patients with longer disease duration at initial examination (6 to 15 years). This may explain why we did not find a strong effect for *GBA*, and is supported by analysis of *GBA* in patients earlier in disease stage^44^. In addition, we only had coverage of the p.E326K variant which is a ‘milder’ mutation^39,45^. Our unbiased genome-wide search suggests that, in addition to *GBA*, there are potentially other genes that are important for PD progression.

Our targeted analysis showed that only a few PD risk variants were nominally associated with progression, similar to the previous PD progression GWAS^24,46^. This suggests that there is minimal overlap in the genetic architecture of PD risk and PD progression. Similarly, the age at onset GWAS showed only a partial overlap with the genetics of PD risk^26^. Given our current sample size, we would have at least 80% power to detect an individual genotype effect which explains 1.8% of the variance in progression. It is now becoming increasingly important to study disease phenotypes in GWASs, as these improve our understanding of the biology of progression.

We did not replicate the finding for the *SLC44A1* variant that was associated with progression to Hoehn and Yahr stage 3 in a previous PD progression GWAS^24^. We have used different methods and a different phenotype to analyse PD progression. Further progression GWASs are needed to replicate both sets of results, and other metrics for PD progression could be analysed, such as mortality.

A strength of this study is the analysis of large, more homogeneous cohorts with relatively long follow-up. Overall, the cohorts were similar although the PPMI patients were younger and earlier in disease stage. One limitation is that longer follow-up is needed to detect variants which may influence progression in later disease stages, such as *GBA*. We may also be underpowered to detect variants with smaller effects on progression. Although the HD GWAS identified significant signals in smaller samples^7^, analysis of PD progression is more complex due to slower progression, greater heterogeneity in genetic risk and rate of progression between patients, and greater dissociation between motor and cognitive progression.

Secondly, our analysis of motor progression may be affected by medication. We used the ‘off’ state motor assessment from PPMI, because patients were medication-naïve at baseline, however in the other cohorts the majority of patients were assessed in their ‘on’ medication state. We did not adjust for levodopa dosage as this is likely to be correlated with progression.

A third limitation is the potential inclusion of patients that have non-PD conditions. We did not find that our results changed substantially when we excluded patients with diagnostic certainty <90%. However certainty data was not available for PPMI, and abnormal dopamine transporter scans cannot differentiate between PD and other degenerative parkinsonian conditions^48^. Despite this, our sensitivity analyses suggest that our results are not being driven by non-PD conditions. Our GWASs also did not identify loci that are associated with PSP risk, including *MAPT, MOBP^49^*, or the variant rs2242367 near *LRRK2* associated with PSP progression^50^.

Many of our top variants had weaker signals when we excluded the 10% fastest and slowest progressing patients, likely due to loss of statistical power. With our duration of follow-up, we are likely to have excluded the majority of non-PD patients as diagnostic accuracy improves after 5 years of disease duration^1,51^ however it is possible that some have not been excluded. Analysis of pathologically-confirmed PD cases is needed to resolve this issue.

This study is the first to use a PCA data reduction method to assess PD progression, based on a successful approach in HD. We robustly replicated the association between *APOE* ε4 and cognitive progression, and have identified other genes which may be associated with progression. These advances are essential to understand the biology of disease progression and nominate therapeutic targets to stop or slow PD progression.

## Data Availability

Anonymised data from Tracking Parkinson’s and Oxford Discovery are available to researchers on application. Please apply via the project coordinators ( respectively). The PPMI data is publicly available on application (https://www.ppmi-info.org/access-data-specimens/download-data/).
Code is available at https://github.com/huw-morris-lab/PD-PCA-progression-GWAS.

https://github.com/huw-morris-lab/PD-PCA-progression-GWAS

## Abbreviations

FUMA: Functional Mapping and Annotation of GWAS
HD: Huntington’s Disease
GWAS: Genome-wide Association Study
LD: Linkage Disequilibrium
MDS-UPDRS: Movement Disorder Society Unified Parkinson’s Disease Rating Scale
MoCA: Montreal Cognitive Assessment
PCA: Principal Components Analysis
PD: Parkinson’s Disease
PPMI: Parkinson’s Progression Markers Initiative
SNP: Single Nucleotide Polymorphism
SD: Standard Deviation

## Data Availability

Anonymised data from Tracking Parkinson’s and Oxford Discovery are available to researchers on application. Please apply via the project coordinators ( respectively). The PPMI data is publicly available on application (https://www.ppmi-info.org/access-data-specimens/download-data/).

Code is available at https://github.com/huw-morris-lab/PD-PCA-progression-GWAS.

## Author Contributions

**1. Research project: A. Conception, B. Organization, C. Execution;**

**2. Statistical Analysis: A. Design, B. Execution, C. Review and Critique;**

**3. Manuscript Preparation: A. Writing of the first draft, B. Review and Critique**.

M.M.X.T.: 2A, 2B, 2C, 3A, 3B

M.A.L.: 1B, 2C, 3B

E.J.: 2C, 3B

R.H.R.: 2C, 3B

H.I.: 2C, 3B

C.B.: 2C, 3B

S.K.: 1B, 3B

M.P.: 2C, 3B

L.H.: 1C

N.M.: 1C, 3B

K.A.G.: 1A, 1B, 1C, 3B

S.L.M.: 1C

N.B.: 1A, 1B, 1C

R.A.B.: 1A, 1B, 3B

D.J.B.: 1A, 1B, 3B

C.B.: 1C

T.F.: 1A, 1B, 3B

J.H.: 1A, 3B

N.W.: 1A, 1B

C.H.W-G.: 1B, 1C, 2C, 3B

M.A.N.: 2C, 3B

A.B.S.: 3B

N.W.W.: 1A, 1B, 2C, 3B

Y.B-S.: 1A, 1B, 2C, 3B

M.T.M.H.: 1A, 1B, 1C, 3B

D.G.G.: 1A, 1B, 1C, 3B

M.S.: 2A, 2C, 3B

H.R.M.: 1A, 1B, 2A, 2C, 3B

## Acknowledgements

Both Tracking Parkinson’s and Oxford Discovery are primarily funded and supported by Parkinson’s UK. Both studies are supported by the National Institute for Health Research (NIHR) Dementias and Neurodegenerative Diseases Research Network (DeNDRoN). Oxford Discovery is also supported by the NIHR Oxford Biomedical Research Centre based at Oxford University Hospitals NHS Trust, and University of Oxford.

This research was supported by the National Institute for Health Research University College London Hospitals Biomedical Research Centre.

This research was supported in part by the Intramural research Program of the NIH, National institute on Aging.

The UCL Movement Disorders Centre is supported by the Edmond J. Safra Philanthropic Foundation.

Data used in the preparation of this article were obtained from the Parkinson’s Progression Markers Initiative (PPMI) database. For up-to-date information on the study, visit www.ppmi-info.org.

PPMI – a public-private partnership – is funded by the Michael J. Fox Foundation for Parkinson’s Research and funding partners (listed in https://www.ppmi-info.org/about-ppmi/who-we-are/study-sponsors/).

## Financial Disclosures for the preceding 12 months

M.M.X.T. is supported Parkinson’s UK. M.A.L. is supported by Parkinson’s UK. E.J. is supported by the Medical Research Council UK. R.H.R. supported through the award of a Leonard Wolfson Doctoral Training Fellowship. N.B. has received payment for advisory board attendance from UCB, Teva Lundbeck, Britannia, GSK, Boehringer and honoraria from UCB Pharma, GE Healthcare, Lily Pharma, Medtronic. He has received research grant support from GE Healthcare, Wellcome Trust, Medical Research Council, Parkinson’s UK and National Institute for Health Research. R.A.B. has received grants from Parkinson’s UK, NIHR, Cure Parkinson’s Trust, Evelyn Trust, Rosetrees Trust, MRC, Wellcome Trust, and EU along with payment for advisory board work from Oxford Biomedica, Living Cell Technologies, Fujifilm Cellular Dynamics Inc, Nova Nordisk, BlueRock Therapeutics, Sana Biotherapeutics, Aspen Neuroscience and UCB along with honoraria from Wiley and Springer for books and editorial work.D.J.B. has received grants from NIHR, Wellcome Trust, GlaxoSmithKline Ltd, Parkinson’s UK and Michael J Fox Foundation. T.F. has received grants from Michael J Fox Foundation, Cure Parkinson’s Trust, Brain Research trust, John Black Charitable Foundation, Rosetrees trust and honoraria for speaking at meetings from Bial, Profile Pharma and Medtronic. N.W.W. is supported by the MRC and NIHR UCLH Biomedical research centre. C.H.W-G is supported by a RCUK/UKRI Research Innovation Fellowship awarded by the Medical Research Council (MR/R007446/1), by the NIHR Cambridge Biomedical Research Centre Dementia and Neurodegeneration Theme (Grant Reference Number 146281), and by the Cambridge Centre for Parkinson-Plus. M.A.N. reports that this work was done under a consulting contract with National Institutes of Health, he also consults for Lysosomal Therapeutics Inc, Neuron23, and Illumina. J.H. is supported by the UK Dementia Research Institute which receives its funding from DRI Ltd, funded by the UK Medical Research Council, Alzheimer’s Society and Alzheimer’s Research UK. He is also supported by the MRC, Wellcome Trust, Dolby Family Fund, National Institute for Health Research University College London Hospitals Biomedical Research Centre. Y.B-S. has received grant funding from the MRC, NIHR, Parkinson’s UK, NIH and ESRC. N.M.W. is supported by Parkinson’s UK. M.T.H. receives grants from Parkinson’s UK, Oxford NIHR Biomedical Research Centre, and MJFF and is an adviser to the Roche Prodromal Advisory and Biogen Digital advisory boards. D.G.G. has received grants from Michael’s Movers, The Neurosciences Foundation, and Parkinson’s UK, and honoraria from UCB Pharma and GE Healthcare, and consultancy fees from Acorda Therapeutics. H.R.M. is supported by the PSP Association, CBD Solutions, Drake Foundation, the Medical Research Council UK, Parkinson’s UK, and Cure Parkinson’s Trust. All other authors did not declare any funding sources that directly contributed to this study. M.M.X.T. takes responsibility for the integrity of the data and the accuracy of the data analysis.

